# Fluctuating High Throughput Serological Assay Results in Recurrent Convalescent Plasma Donors

**DOI:** 10.1101/2020.10.25.20219147

**Authors:** Larry L. Luchsinger, Shiraz Rehmani, Andrew Opalka, Donna Strauss, Christopher D. Hillyer, Patricia Shi, Bruce S. Sachais

**Author notes:** Address correspondence to Bruce Sachais and Larry Luchsinger, and.

## Abstract

The clinical and scientific communities rely on serology testing to analyze the degree of antibody-mediated immunity afforded to recovered patients from SARS-CoV-2 infection. Neutralizing antibodies present in COVID-19 convalescent plasma (CCP) remains a practical therapy to treat COVID-19 patients requiring hospitalization. However, it remains unclear how long antibody levels persist in CCP donors after recovery. An accurate estimation of antibody kinetics in CCP donors provide an important observation to further define the extent of long-term immunity in recovered patient and simultaneously inform CCP collection processes in efforts to improve CCP dosing and therapeutic outcome. In this study, we analyzed 63 donors and measured antibody levels using two high throughput screening assays (HTSA) designed to detect antibodies targeting the spike protein (S1) and nucleocapsid protein (NP) of SARS-CoV-2 and monitored antibody levels between 2-8 consecutive donations. We show that anti-S1 antibody levels, as measured using the Ortho Total Ig HTSA, increased over time in repeat CCP donors while anti-NP antibody levels, as measured using the Abbott IgG HTSA, were unchanged or decreased over time. When we normalized these data, we found that both the absolute levels of anti-S1 antibodies and the ratio between S1 and NP antibodies tends to increase over time. These data have important implications for the convalescent donation process, patient protection from future infection and characterization of the SARS-CoV-2 immune response.

## Introduction

For over a century, passive antibody transfer using convalescent plasma from recovered patients has been used to accelerate the immune response in morbid patients.(1, 2) Before vaccines were developed, this method has been used to treat outbreaks of several novel infectious diseases including measles, mumps, and influenza.(3) Although vaccines and monoclonal antibody therapies are being developed for novel coronavirus, it will still take time before these therapies are widely available to the population.(4) Therefore, convalescent plasma remains a viable approach to treat coronavirus disease 2019 (COVID-19), which is caused by severe acute respiratory syndrome coronavirus 2 (SARS-CoV-2) infection.(5) In support of this approach, several studies have shown efficacy of COVID-19 convalescent plasma (CCP) in hospitalized patients, particularly when treated early.(6, 7) However, evidence for therapeutic CCP efficacy still requires definitive support from large randomized clinical trials

Central to efficacy of CCP therapy is the accurate selection of hyperimmune plasma units, which require accurate quantification of antiviral antibodies. High throughput serological assays (HTSA), which rely on chemiluminescent immunoassay with high dynamic range, can be introduced in the population at scale using clinical laboratory infrastructure to rapidly measure antibody levels of CCP. As such, HTSAs could potentially be a useful tool to study SARS-CoV-2 and to improve CCP donor screening and antibody titration accuracy of CCP units. Serological assays rely on accurate recognition and ideally quantification of antibodies that recognize viral antigens specific to SARS-CoV-2. Coronaviruses have four major structural proteins; spike protein (S1), nucleocapsid protein (NP), membrane protein, and envelop protein.(8) Research conducted on 2005 SARS-CoV-1 and Middle East respiratory syndrome Coronavirus (MERS-CoV), which are highly related to SARS-CoV-2, found that recovered individuals produced the strongest immunogenic antibodies against antigens of the S- and N-proteins.(9) Thus, the development of serological tests for SARS-CoV-2 antibodies has focused heavily on the detection of antibodies against S1 and NP proteins.

Recently, we have shown that HTSA assays, specifically those that measure anti-S1 antibodies, demonstrated the strongest correlation with neutralizing antibodies, which are critical for mediating the antiviral activity that supports effective convalescent plasma therapy.(10) Herein, we used the Ortho Total Ig and Abbott IgG assays measure anti-S1 and anti-NP antibodies, respectively to investigate antibody levels of recurring CCP donors in the New York City metro area.

## Results

CP donors were evaluated for antibody levels over the duration of 2-8 donations. The time between first and last donation ranged from 14-45 days with a median difference of 28 days (**Figure 1A**). For each donation, plasma antibodies against SARS-CoV-2 were quantified using the Ortho Total Ig and Abbott IgG HTSA assays. Serology values from 63 unique donors using the Ortho HTSA assay showed a statistically significant increase in the median value of 110 AU (95% CI: 76.6 – 151 AU) at the first donation to 226.5 AU (95% CI: 109 – 358 AU) at the last donation (**Figure 1B**). Intriguingly, serology values using the Abbott HTSA assay did not show a significant change, which showed median value of 5.45 AU (95% CI: 4.3 – 6.39 AU) at the first donation to 5.17 AU (95% CI; 4.72 – 5.96) at the last donation (**Figure 1C**). Transformation of these serology data as a ratio between last and first donation demonstrated an average increase of 226% in Ortho HTSA values, while Abbott HTSA values slightly decreased with an average decrease of 4% between repeat donations (**Figure 1D**). These data show that convalescent plasma donors acquire higher amounts of S1 antibodies between 1 and 2 months of their first donation.

**Figure 1:**
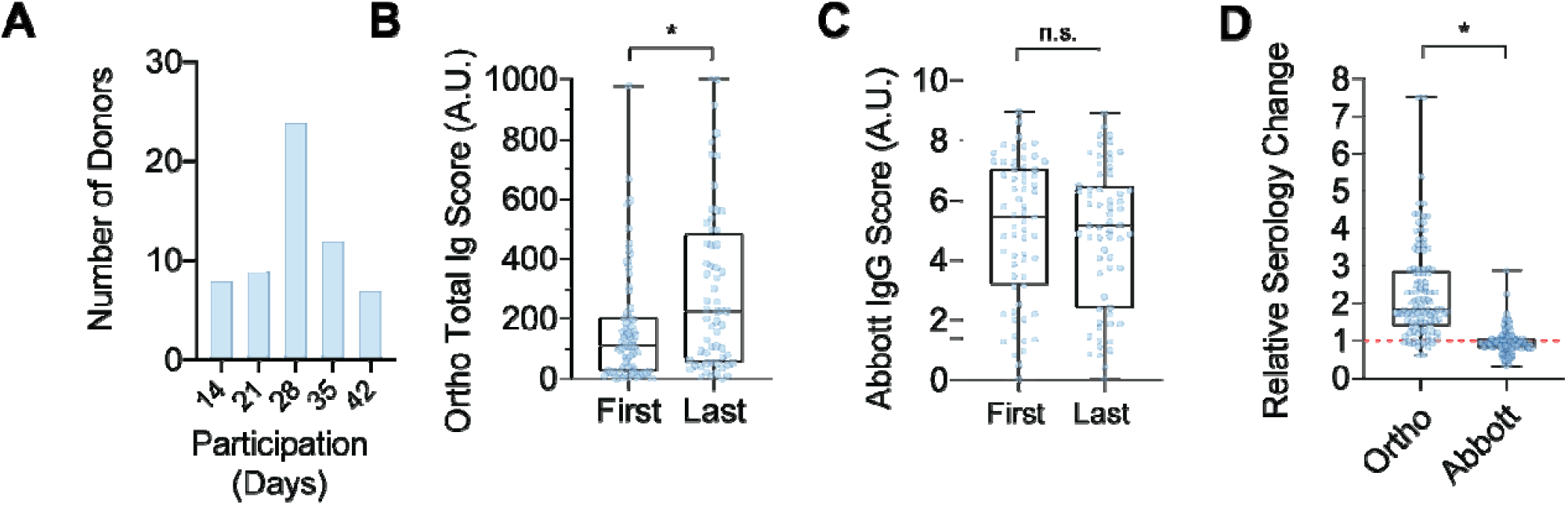
Characterization of repeat donor serology results. **A;** Range of time, in days, between first and last donations from repeat convalescent plasma donors. **B;** Ortho Total Ig HTSA values from repeat convalescent plasma donors at the first and last donation. Box plot represents first quartile, median and third quartile, whiskers represent range of values. N=63; student’s t-test, * p < 0.01. **C;** Abbott IgG HTSA values from repeat convalescent plasma donors at the first and last donation. Box plot represents first quartile, median and third quartile, whiskers represent range of values. N=63; Mann-Whitney test. **D;** Difference in HTSA results from repeat convalescent plasma donors between the first and last donation. Dotted red line represents relative value at first donation. Box plot represents first quartile, median and third quartile, whiskers represent range of values. N=63; Mann-Whitney test, * p < 0.01.

To further characterize the kinetics of antibody levels over time, we plotted the serology results versus the change in time from the first donation for Ortho (**Figure 2A**) and Abbott (**Figure 2B**) HTSAs. Visual examination of the plots shows a trending increase in Ortho values with time as opposed to Abbott values, which appear more ambiguous. To express the change in serology results over time, we fitted each donor series to a linear regression and calculated the slope of the line fit (**Figure 2C**). Interestingly, 96% of repeat donors showed positive slopes, with a median value of 2.4 (95% CI: 1.7 – 4.8, range −2.6 – 19.06) which denotes an increase in Ortho values over time. Conversely, Abbott HTSA values showed inconsistency of slopes between repeat donors, with a median value of −0.01 (95% CI: – 0.02 to 0, range. −0.06 – 0.12), which implies no change in Abbott values over time. We noted that the data showed a variety of goodness-of-fit values (r^2^), which is the measure of variance intrinsic to the linear regression. Thus, we subdivided repeat donor series into various categories of r^2^ values (**Figure 2D**). Interestingly, the mean slope of Ortho scores with time was statistically higher in repeat donor series with less intrinsic variability (r^2^ > 0.9) compared to those with r^2^ < 0.5, while no significant difference was found in the mean slope of Abbott scores between these groups. These data further illustrate that anti-S1 antibodies measured by the Ortho HTSA appear to increase with time in repeat donors while anti-NP antibodies measured by the Abbott IgG HTSA appear to persist or slightly decrease with time.

**Figure 2:**
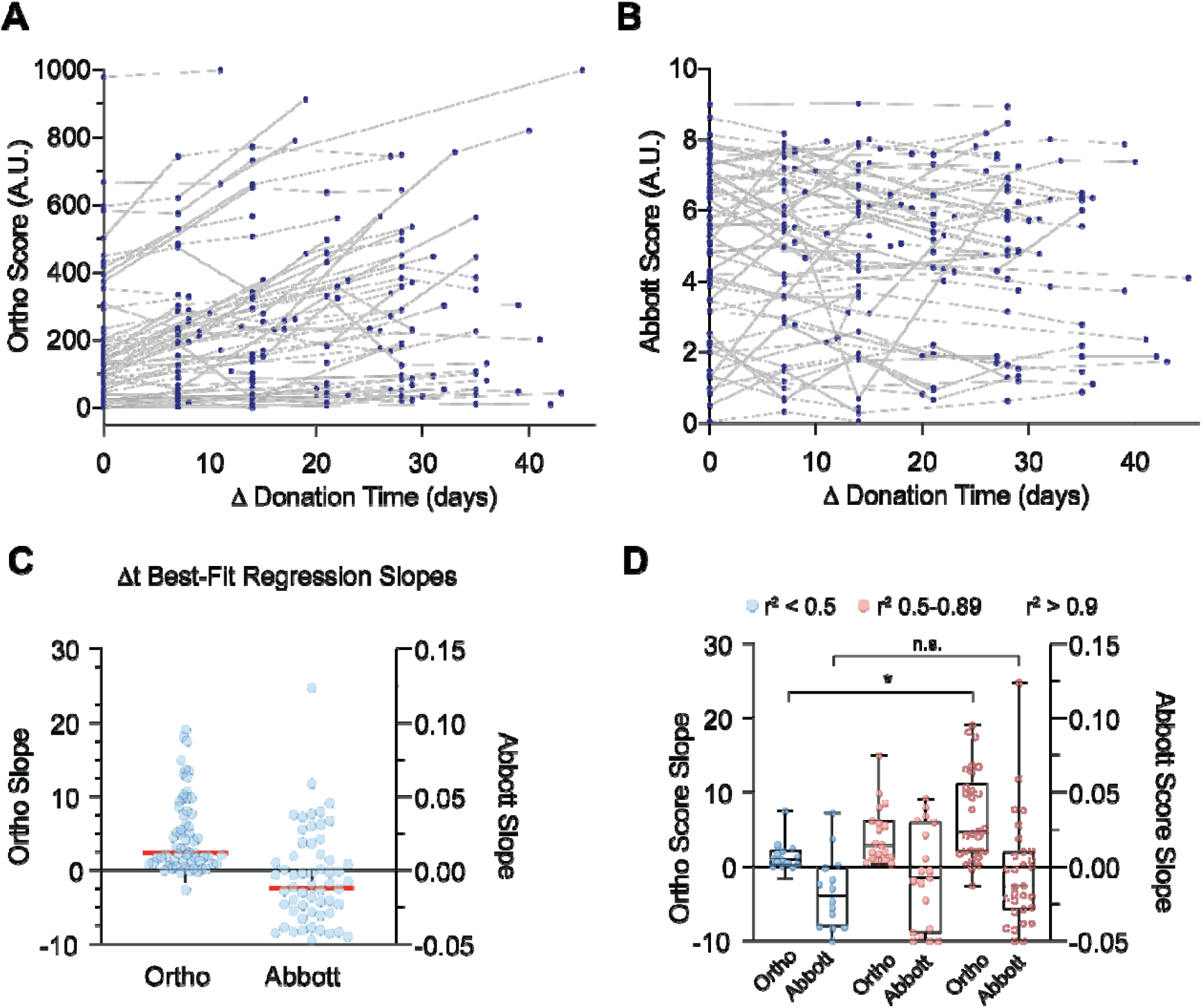
Quantification of repeat donor serology values over time. **A;** Dot plot of consecutive Ortho Total Ig HTSA values from the first to last donations in repeat convalescent plasma donors. **B;** Dot plot of consecutive Abbott IgG HTSA values from the first to last donations in repeat convalescent plasma donors. **C;** Linear regression slope values from repeat convalescent plasma donors. Ortho and Abbott HTSA scales are shown on the left and right ordinates, respectively. Red line indicates median value. **D;** Linear regression slope values from repeat convalescent plasma donors grouped into goodness-of-fit categories as indicated. Box plot represents first quartile, median and third quartile, whiskers represent range of values. N=63; Mann-Whitney test, * p < 0.01.

It is difficult to directly compare serological assays to each other, mostly because the scale and range of assay values vary between platforms and target antigens. Therefore, we normalized these factors by calculating the Ortho Total Ig/Abbott IgG HTSA Quotient (Q, see Equation 1 in Methods). In this way, the relative values between Ortho and Abbott assays (or effectively, anti-S1 and anti-NP antibodies) can be compared more easily between donors. We calculated Q for repeat donor series and plotted these values with time (**Figure 3A**). The results show a large range of Q values between donors at each donation but, as noted of Ortho HTSA values with time (**Figure 2A)**, Q values display a trending increase with time. To further normalize these data between donors, we plotted ΔQ values (see Equation 2 in Materials and Methods), in which the Q value of the first donation was used to normalize subsequent Q values from repeat donors, resulting in a ΔQ for each donor of 1.0 at the first donation (**Figure 3B**). Interestingly, the plot of ΔQ over time showed a clear increase in ΔQ values in almost all repeat donors with time, suggesting the ratio between S1 to NP antibodies increased over time. Quantification of ΔQ values between last and first donation showed a median increase of 2.1 (**Figure 3C**, 95% CI: 1.5 – 2.3, range 0.5 – 9.6). Taken together, these data demonstrate that convalescent plasma donors tend to increase the absolute amount of anti-S1 antibodies and the ratio between anti-S1 to anti-NP antibodies over the first 1-2 months from donation.

**Figure 3:**
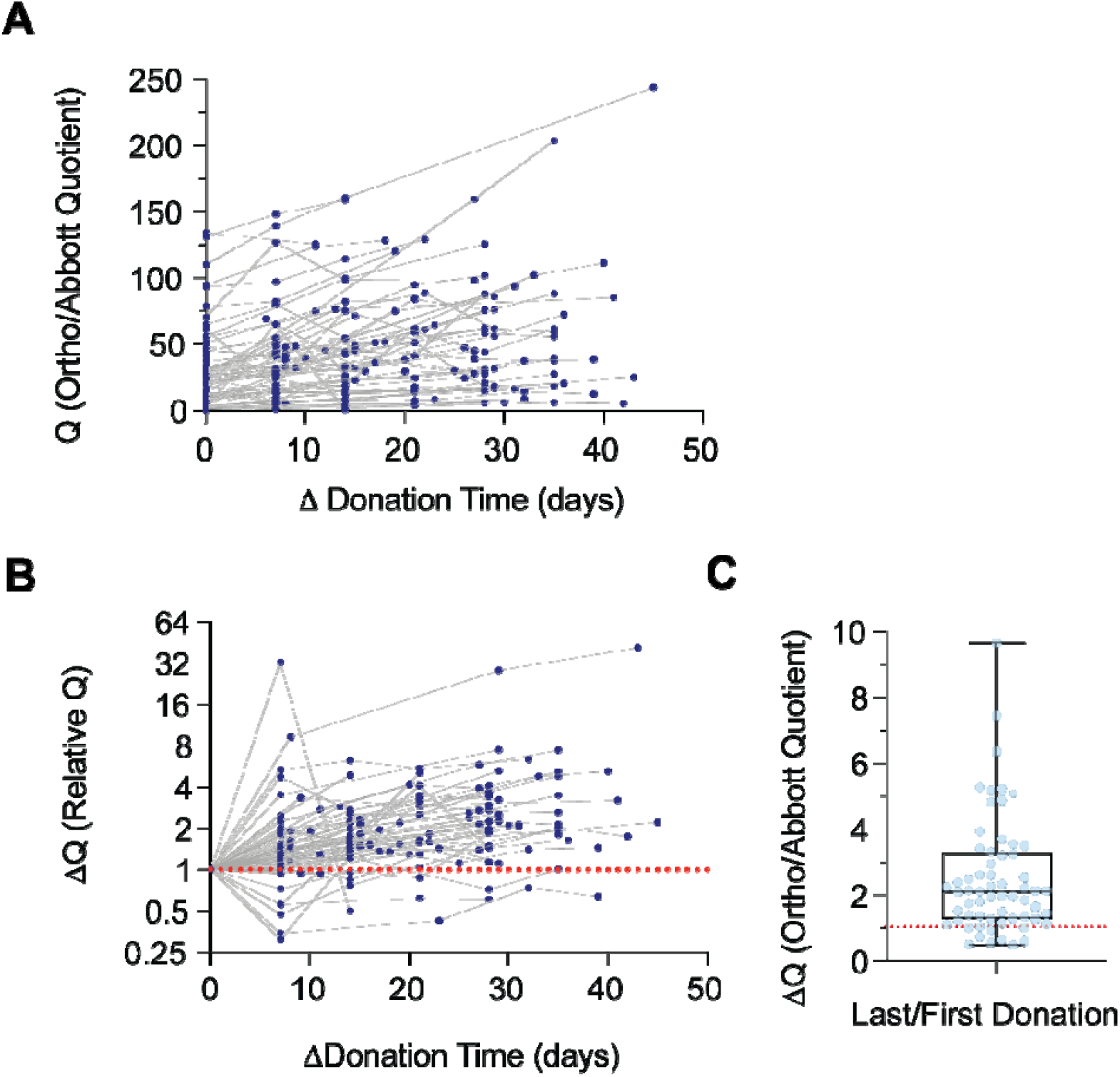
Normalization of repeat donor serology values over time. **A;** Dot plot of the Ortho/Abbott HTSA Quotient (Q) over time from individual repeat convalescent plasma donations. **B;** Q values relative to first donation (ΔQ) with time from individual repeat convalescent plasma donations. Dotted red line indicates value relative to first donation. **C;** Difference in ΔQ values from repeat convalescent plasma donors between the first and last donation. Dotted red line indicates value relative to first donation. Box plot represents first quartile, median and third quartile, whiskers represent range of values.

## Discussion

The value of convalescent plasma as a viable therapy for COVID-19 was quickly recognized by the medical community early in the pandemic. In response, the New York Blood Center launched the first COVID-19 convalescent plasma program in the country, which has distributed over 50,000 units to date.(11) Early in the pandemic, a number of small studies showed evidence favoring effect of reducing mortality in hospitalized COVID-19 patients who received transfusions; however, they generally lacked sufficient analytical power and randomization necessary for definitive evidence.(12) Recently, convalescent plasma therapy has received Emergency Use Authorization from the FDA, following a comprehensive evaluation of CP donor efficacy in over 35,000 transfusions performed in the United States leading up to July of 2020.(13) While this study found a wide deviation in the odds ratios of CP effectiveness, it became the first study to incorporate an HTSA assay as a means to titrate antibody levels of CP units, which the authors used to stratify the outcome of the CP therapy. Although a large variation was still observed, likely due to differences in patient morbidity and duration of infection at the time of transfusion, the authors found that CP units with high Ortho IgG HTSA scores showed improved survival odds compared to patients that received low titer CP units. As such, HTSA data can potentially be used to address the question as to effective dosing of convalescent plasma.(14) Since neutralizing antibodies are thought to be the essential functional unit of CP, and as anti-S1 antibodies highly correlate with antiviral neutralizing activity,(15) HTSAs that correlate with neutralizing activity offer a rational approach to grade the relative “potency” of CCP units to help standardize the clinical dosing. Although there currently remains no defined threshold of neutralizing antibody necessary to inhibit COVID-19, CCP units likely include various soluble factors that could be beneficial to recipients, such as cytokines, platelets or soluble factors.(16) Thus, an effective titration strategy of CCP using HTSA assays could provide a more apportionable therapeutic with defined serology characteristics and supportive soluble factors to aid the recovery of recipients.

The kinetics of antibody development are an important question in our understanding of recovery from SARS-CoV-2. In this study, we found anti-S1 antibodies appeared to increase approximately 2-fold in repeat CP donors between 14 and 45 days from first donation. This is an important observation for several reasons. First, the current convalescent plasma program criterion does not require antibody screening of candidates before their first donation. From our evidence, CCP collection from repeat donors with an appreciable amount of antibody should be encouraged to continue to donate so long as anti-S1 antibodies are consistent or increasing with time, as measured by HTSA assays, relative to their first donation. Second, our finding that a net increase in anti-spike antibodies in convalescent plasma donors occurs with time offers some insight into the duration of immunity afforded to recovering patients of COVID-19 if, in fact, antiviral antibodies are important to preventing reinfection. Third, measurement of antibody kinetics of multiple epitopes may be useful for epidemiological and immunological study of COVID-19. Given that vaccination strategies hinge on this effect, observing the natural increasing levels of anti-S1 antibodies is likely a natural immune response, which may be similar or different in response to vaccination.

While our study provides an intriguing analysis of antibody responses over time in convalescent plasma donors, we recognize some limitations. First, our sample size was considerable but not large enough to make generalizations about specific demographics. Further, our longitudinal analysis of recurring CP donors is limited to 45 days from the first donation. Therefore, future studies designed to monitor antibody levels over a longer time period in different age groups, ethnic backgrounds and between genders for extended periods of time are required. Finally, our longitudinal analysis of serological results are somewhat consistent with the recent report by Muechsch *et al*. where the authors used a variety of HTSA assays.(17) Indeed, both of our studies included that the Abbott IgG assay showed a decline in anti-NP antibodies with time and some anti-S1 assays showed increase over time. However, Muechsch *et al*. concluded that neutralizing activity in a random group of blood donors decreased between 20-80 days post-diagnosis. However, the minimal level of neutralizing activity or serology value needed to confer effective CP therapy in transfusion recipients or serum antibody levels to prevent reinfection remains to be determined.

## Data Availability

All raw data is available upon request to the corresponding authors.

## Declarations

### Ethics approval and consent to participate

Approval for donation and collection of blood from donors was attained by written consent. All donors were over 16 years of age. Ethical oversight of seroprevalence donation protocols was obtained from the institutional review board of the New York Blood Center.

### Availability of data and materials

All raw data presented in this manuscript is available on request from the corresponding author.

### Authors’ Contributions

AO, DS and BS designed the study and collected the data. LL, AO and SR performed data analysis and statistical calculations. LL, BS, DS and CDH co-wrote the manuscript. All authors read and approved the submission of the manuscript.

## Acknowledgements

We thank Jill Alberigo, Amanda Brites and Kelly Brightman and the laboratory staff from the Rhode Island Blood Center for their help with performing the Ortho Anti-SARS-CoV-2 Total Ig Test and the Abbott Anti-SARS-CoV-2 IgG Test, managing donation samples and relaying test results.

## Methods

### CCP Donors and Sample Preparation

From April 2020 – July 2020, consecutive donations from CP donors (n=63) completed a convalescent plasma donation. Donors were qualified for CCP if they had documented SARS-CoV2, had recovered and were symptom free for a minimum of 14 days, and met all allogeneic blood donation criteria. Donors were allowed to donate every 7 days, with up to 8 donations in the first three months. Thereafter, donors could donate CCP following the standard 28-day donation interval for plasma as long as appropriate anti-SARS-CoV2 antibodies continued to demonstrate. Plasma samples were extracted, aliquoted to minimize freeze-thaw cycles, and stored at −80°C. Donor blood samples were tested using the Ortho VITROS™ SARS-CoV-2 Total Ig assay and the Abbott SARS-CoV-2 IgG assay.

### High-throughput Serology Assays

Samples were analyzed using the Abbott SARS-CoV-2 IgG chemiluminescent microparticle immunoassay using the Abbott Architect i2000SR (Abbott Core Laboratories), as well as the VITROS Immunodiagnostic Products Anti-SARS-CoV-2 Total Test using the VITROS 5600 (Ortho Clinical Diagnostics). All assays were performed by trained RIBC employees according to the respective manufacturer standard procedures.

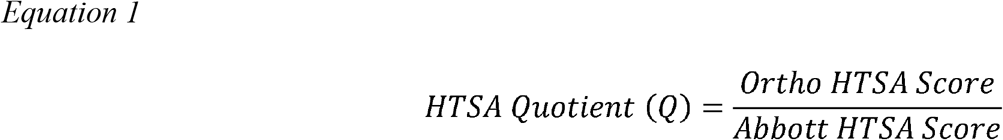

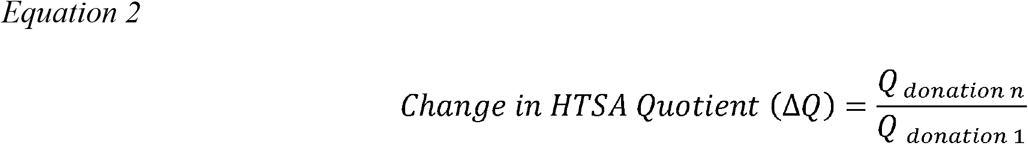

## References

1. Cunningham AC, Goh HP, Koh D. Treatment of COVID-19: old tricks for new challenges. Crit Care. 2020;24(1):91.

2. Selvi V. Convalescent Plasma: A Challenging Tool to Treat COVID-19 Patients—A Lesson from the Past and New Perspectives. BioMed Research International. 2020;2020:2606058.

3. Casadevall A, Pirofski LA. The convalescent sera option for containing COVID-19. J Clin Invest. 2020;130(4):1545–8.

4. Cai X, Ren M, Chen F, Li L, Lei H, Wang X. Blood transfusion during the COVID-19 outbreak. Blood Transfus. 2020;18(2):79–82.

5. Zhou G, Zhao Q. Perspectives on therapeutic neutralizing antibodies against the Novel Coronavirus SARS-CoV-2. Int J Biol Sci. 2020;16(10):1718–23.

6. Liu STH, Lin HM, Baine I, Wajnberg A, Gumprecht JP, Rahman F, et al. Convalescent plasma treatment of severe COVID-19: a propensity score-matched control study. Nat Med. 2020.

7. Salazar E, Christensen PA, Graviss EA, Nguyen DT, Castillo B, Chen J, et al. Treatment of Coronavirus Disease 2019 Patients with Convalescent Plasma Reveals a Signal of Significantly Decreased Mortality. Am J Pathol. 2020;190(11):2290–303.

8. Zhong X, Yang H, Guo ZF, Sin WY, Chen W, Xu J, et al. B-cell responses in patients who have recovered from severe acute respiratory syndrome target a dominant site in the S2 domain of the surface spike glycoprotein. J Virol. 2005;79(6):3401–8.

9. Tai W, He L, Zhang X, Pu J, Voronin D, Jiang S, et al. Characterization of the receptor-binding domain (RBD) of 2019 novel coronavirus: implication for development of RBD protein as a viral attachment inhibitor and vaccine. Cell Mol Immunol. 2020.

10. Luchsinger LL, Ransegnola B, Jin D, Muecksch F, Weisblum Y, Bao W, et al. Serological Assays Estimate Highly Variable SARS-CoV-2 Neutralizing Antibody Activity in Recovered COVID19 Patients. J Clin Microbiol. 2020.

11. Budhai A, Wu AA, Hall L, Strauss D, Paradiso S, Alberigo J, et al. How did we rapidly implement a convalescent plasma program? Transfusion. 2020.

12. Joyner MJ, Klassen SA, Senefeld J, Johnson PW, Carter RE, Wiggins CC, et al. Evidence favouring the efficacy of convalescent plasma for COVID-19 therapy. medRxiv. 2020:2020.07.29.20162917.

13. Joyner MJ, Senefeld JW, Klassen SA, Mills JR, Johnson PW, Theel ES, et al. Effect of Convalescent Plasma on Mortality among Hospitalized Patients with COVID-19: Initial Three-Month Experience. medRxiv. 2020:2020.08.12.20169359.

14. Al-Riyami AZ, Schafer R, van den Berg K, Bloch EM, Estcourt LJ, Goel R, et al. Clinical use of Convalescent Plasma in the COVID-19 pandemic: a transfusion-focussed gap analysis with recommendations for future research priorities. Vox Sang. 2020.

15. Wang C, Li W, Drabek D, Okba NMA, van Haperen R, Osterhaus A, et al. A human monoclonal antibody blocking SARS-CoV-2 infection. Nat Commun. 2020;11(1):2251.

16. Daniele F, Marco T, Guido A, Fabrizio M. What is the optimal usage of Covid-19 convalescent plasma donations? Clin Microbiol Infect. 2020.

17. Muecksch F, Wise H, Batchelor B, Squires M, Semple E, Richardson C, et al. Longitudinal analysis of clinical serology assay performance and neutralising antibody levels in COVID19 convalescents. medRxiv. 2020:2020.08.05.20169128.

